# Effectiveness of therapeutic exercise in patients with fibromyalgia on psychosocial and clinical outcomes: a systematic review protocol

**DOI:** 10.1101/2025.04.27.25326100

**Authors:** Giuseppe Enea, Angelo Ginestra, Rosario Fiolo, Filippo Brighina, Francesco Piampiano, Claudio Tarantino, Elena Notararigo, Gabriele Assanto

## Abstract

**Background:** Fibromyalgia (FMS) is a chronic syndrome characterized by widespread musculoskeletal pain, fatigue, sleep disturbances, and psychological symptoms such as anxiety and depression (1). Therapeutic exercise is recommended as a non-pharmacological treatment for FMS management; however, its effectiveness on clinical and psychosocial parameters remains uncertain (2). This systematic review aims to synthesize the available evidence on the impact of therapeutic exercise in patients with FMS, assessing both clinical outcomes (pain, physical function) and psychosocial outcomes (quality of life, psychological well-being).

**Methods:** This review will include randomized controlled trials (RCTs) published between 2014 and 2024 that evaluate the effectiveness of therapeutic exercise in adult patients with FMS. A comprehensive literature search will be conducted in PubMed, PEDro, Embase, and Cochrane CENTRAL using predefined search strategies. Study selection will follow a blinded process conducted by two independent reviewers, with any disagreements resolved by a third reviewer. Data will be extracted and synthesized into tables and descriptive summaries, without the use of inferential statistical methods or meta-analysis.

**Ethics and Dissemination:** As this is a systematic review, ethical approval is not required. The results will be disseminated through publications in peer-reviewed journals and presentations at national and international scientific conferences, with the aim of supporting clinical practice and research in FMS management.

## 1. Introduction

Fibromyalgia (FMS) is a complex chronic syndrome characterized by widespread musculoskeletal pain, persistent fatigue, insomnia, stiffness, and cognitive impairments. It is often associated with psychological conditions such as depression and anxiety (1). FMS affects approximately 2–4% of the general population, with a higher prevalence in women than in men. In Italy, an estimated 1.5–2 million people are affected by FMS, although its diagnosis is often hindered by a lack of awareness and the variability of symptoms among patients (3).

The etiology of FMS remains unclear; however, genetic predisposition, traumatic events, and alterations in pain modulation are believed to contribute to its development (4). While chronic pain is the predominant symptom, other manifestations, such as fatigue and sleep disturbances, can have an even greater impact on patients’ quality of life.

The therapeutic approach to FMS is multidimensional, encompassing pharmacological treatments, educational interventions, psychological support, and physical therapies. However, pharmacological therapies are effective in only about 30% of patients, highlighting the crucial role of non-pharmacological approaches such as therapeutic exercise. This intervention is recommended by clinical guidelines due to its potential to reduce pain, improve physical function, and enhance psychological well-being. Nevertheless, the optimal type and intensity of exercise remain uncertain, emphasizing the need for further systematic syntheses of the available evidence.

This protocol aims to evaluate the effectiveness of therapeutic exercise in the management of FMS, focusing on clinical outcomes (e.g., pain, physical function) and psychosocial outcomes (e.g., quality of life, psychological well-being).

## 2. Objective

The aim of this systematic review is to analyze and synthesize the evidence on the impact of therapeutic exercise in patients with fibromyalgia. The outcomes of interest include:

- **Clinical outcomes:** pain, physical function, and overall health status.
- **Psychosocial outcomes:** quality of life, psychological well-being, anxiety, and depression levels.

## 3. Methodology

### 3.1 Research Question (PICO)

- **Population (P):** Adults (≥18 years) with a clinical diagnosis of fibromyalgia according to the American College of Rheumatology (ACR) criteria.
- **Intervention (I):** Therapeutic exercise programs, including aerobic exercise, resistance training and stretching.
- **Comparator (C):** No intervention, standard care, or alternative treatments.
- **Outcome (O):** Improvements in pain, physical function, quality of life, psychological well-being, and other psychosocial parameters.

### 3.2 Inclusion Criteria

The review will include:

- Randomized controlled trials (RCTs) published between January 2014 and June 2024.
- Studies published in English.
- Adult participants (≥18 years) diagnosed with fibromyalgia.
- Interventions incorporating therapeutic exercise as the primary treatment or in combination with other non-pharmacological therapies.

### 3.3 Exclusion Criteria

The review will exclude:

- Studies exclusively evaluating pharmacological therapies or alternative treatments such as yoga, acupuncture, Reiki, Tai Chi, Qi Gong, and Pilates.
- Observational studies, case reports, narrative reviews, or non-systematic reviews.

### 3.4 Information Sources

The literature search will be conducted in the following electronic databases:

1. **PubMed**
2. **PEDro**
3. **Embase**
4. **Cochrane Central Register of Controlled Trials (CENTRAL)**

### 3.5 Search Strategy

Search strategies will be tailored to each database and will include combinations of keywords such as “fibromyalgia” “therapeutic exercise” “aerobic exercise” “pain management” “quality of life” and related terms. Temporal filters (2014–2024) and language restrictions (English) will be applied to optimize search results.

### 3.6 Article Screening

Following the extraction of articles from online databases, a two-stage selection process will be conducted:

1. **Title and Abstract Screening:** Two independent reviewers (EN and GA) will assess titles and abstracts to identify relevant studies based on the inclusion and exclusion criteria. Articles deemed potentially eligible will proceed to the next phase.
2. **Full-Text Review:** Selected articles will be analyzed in full to verify their eligibility. Any discrepancies between the two reviewers will be resolved by a third reviewer (AG).

To ensure transparency and reproducibility, the screening process will be documented according to the PRISMA flow diagram, indicating the number of studies identified, excluded, and included in the final synthesis.

### 3.7 Data Analysis

Selected studies will undergo data extraction, including participant characteristics, intervention details, reported outcomes, follow-up duration, and key results (GA, EN, GE). The methodological quality of the studies will be assessed using the PEDro scale, and results will be synthesized qualitatively, considering data heterogeneity.

### 3.8 Statistical Analysis

Data analysis will be conducted using descriptive methods, summarizing study characteristics and key outcomes in tables and summary reports.

Results will be organized in comparative tables, highlighting study design, sample size, population characteristics, intervention type, follow-up duration, and main outcomes. Additionally, key efficacy results for each study will be reported, with a qualitative synthesis of the available evidence.

No meta-analysis or inferential statistical analyses will be performed. Instead, the objective is to provide a structured and comparative overview of the analyzed studies to identify emerging trends and formulate evidence-based recommendations.

## 4. Expected Results

This study aims to identify the most effective therapeutic exercise programs for improving pain, physical function, and psychological well-being in patients with fibromyalgia. The results will contribute to:

1. Clarifying the most appropriate exercise modalities for patients with FMS.
2. Providing practical recommendations for clinicians on the personalization of rehabilitation interventions.
3. Strengthening the inclusion of therapeutic exercise in FMS management guidelines.

## Data Availability

All data produced in the present study are available upon reasonable request to the authors

## Ethics and Dissemination

This study is a systematic review of the literature and does not involve direct participation of patients or human subjects. Therefore, approval from an Ethics Committee is not required.

The results will be disseminated through publications in peer-reviewed scientific journals and presentations at national and international conferences. Additionally, they will be shared with healthcare professionals and researchers to support the improvement of evidence-based fibromyalgia management strategies.

## Author Contributions

GE, GA, and EN conceptualized and designed the study protocol.

GE, GA, EN, AG, FP, and CT contributed to defining the study objectives, selecting the inclusion criteria, and planning data extraction.

All authors reviewed and approved the final version of the protocol.

